# Deep Learning Approaches to Identify Patients within the Thrombolytic Treatment Window

**DOI:** 10.1101/2022.01.26.22269260

**Authors:** Jennifer S Polson, Haoyue Zhang, Kambiz Nael, Noriko Salamon, Bryan Y Yoo, Suzie El-Saden, Sidney Starkman, Namkug Kim, Dong-Wha Kang, William F Speier, Corey W Arnold

## Abstract

1.

**Background:** Treatment of acute ischemic stroke is heavily contingent upon time, as there is a strong relationship between time clock and tissue progression. We sought to a develop a deep learning algorithm for classifying time since stroke (TSS) from MR images by comparison to neuroradiologist assessments of imaging signal mismatch and evaluation on external data.

**Methods:** This retrospective study involved patients who underwent MRI from 2011-2019. Models were trained to classify TSS within 4.5 hours; performance metrics with confidence intervals were reported on both internal and external evaluation sets.

**Results:** A total of 772 patients (66 ± 9 years, 319 women) were used for model development and evaluation. Three board-certified neuroradiologists’ assessments, based on majority vote, yielded a sensitivity of 0.62, a specificity of 0.86, and a Fleiss’ kappa of 0.46. The deep learning method performed similarly to radiologists and outperformed previously reported methods, with the best model achieving an average evaluation accuracy, sensitivity, and specificity of 0.726, 0.712, and 0.741, on an internal cohort and 0.724, 0.757, and 0.679, respectively, on an external, unseen evaluation cohort from another institution.

**Conclusion:** This model achieved higher generalization performance on external evaluation datasets than the current state of the art for TSS classification.

## 2. Introduction

For acute ischemic stroke (AIS) patients, the benefit of thrombolytic therapy is positively associated with earlier reperfusion time.^1,2^ Until recently, thrombolysis was only recommended for AIS patients with a known symptom onset time (TSS) within 4.5 hours.^2,3^ AIS with unknown or unclear TSS has been reported in as many as 35% of patients.^4^ In one study only 6.5% of patients hospitalized for AIS received intravenous thrombolysis, with unknown TSS being the primary reason for treatment exclusion.^5^ Many studies have sought clinical factors to assess eligibility and risk for thrombolytics, with significant focus on neuroimaging.^6–8^ The Efficacy and Safety of MRI-Based Thrombolysis in Wake-Up Stroke (WAKEUP) trial showed that signal mismatch between diffusion-weighted imaging (DWI) and fluid-attenuated inversion recovery (FLAIR) mismatch can be used to select AIS patients with unknown TSS for thrombolytic treatment.^9^ Accordingly, use of DWI-FLAIR mismatch is now recommended (level IIa) to identify unwitnessed AIS patients who may benefit from thrombolytic treatment in the updated American Heart Association-American Stroke Association (AHA-ASA) guidelines.^2^ DWI-FLAIR mismatch, like any subjective assessment, is prone to reader variability that may result in erroneous exclusion of patients who could benefit from thrombolytic treatment.^10^ TSS on the other hand, is an objective surrogate biomarker in clinical settings. Thus, an automated method that classifies TSS could broaden the number of patients eligible for thrombolytic treatment. Machine learning has shown utility for stroke-specific clinical decision support.^11,12^ Deep learning specifically has been widely explored for imaging-based tasks.^13,14^ However, models may suffer from reduced performance on unseen external datasets, requiring external evaluation of these algorithms.^15^

In this retrospective work, we evaluate three methods to assess TSS: DWI-FLAIR mismatch assessments by neuroradiologists, a previously published state-of-the-art machine learning method, and our new deep learning algorithm. We report their performance and compare them to the mismatch assessments. Using an external evaluation set, we explore the algorithms’ generalizability by varying the amounts of new data used for model refinement and retraining. Finally, we use occlusion and gradient-based visualizations to gain insight into model behavior.

## 3. Materials and Methods

### 3.1. Datasets

This study was retrospective, observational dataset comprising patients from two institutions. Patients were included in the cohorts based on the following inclusion criteria: 1) diagnosis with acute ischemic stroke, 2) received pretreatment MRI protocol with DWI, FLAIR and apparent diffusion coefficient (ADC) series without motion degradation, and 3) known TSS within 24 hours of image acquisition. The internal cohort comprised 417 patients treated from 2011-2019; the patient workflow is summarized in Figure 1. The second dataset, published by Lee et al., totaled 355 patients, with more extensive exclusion criteria previously described.^12^ To ensure consistency across both datasets, images were subjected to a preprocessing pipeline.^16^ Images had the neck and skull removed via the Brain Extraction Tool and underwent N4-bias field correction.^17,18^ The T2 series was registered to the MNI-152 T2 atlas, then served as the fixed volume for co-registration. Finally, they were subjected to z-score intensity normalization and histogram matching.^19^ Patient were binarized into two groups: a positive label was given to those who underwent imaging within 4.5 hours of known symptom onset, and a negative label assigned to patients who underwent imaging outside this window. The two datasets were divided into development and evaluation sets to be used for training and testing following an 80/20 random stratified split with respect to the target label as well as clinical parameters.

**Figure 1:**
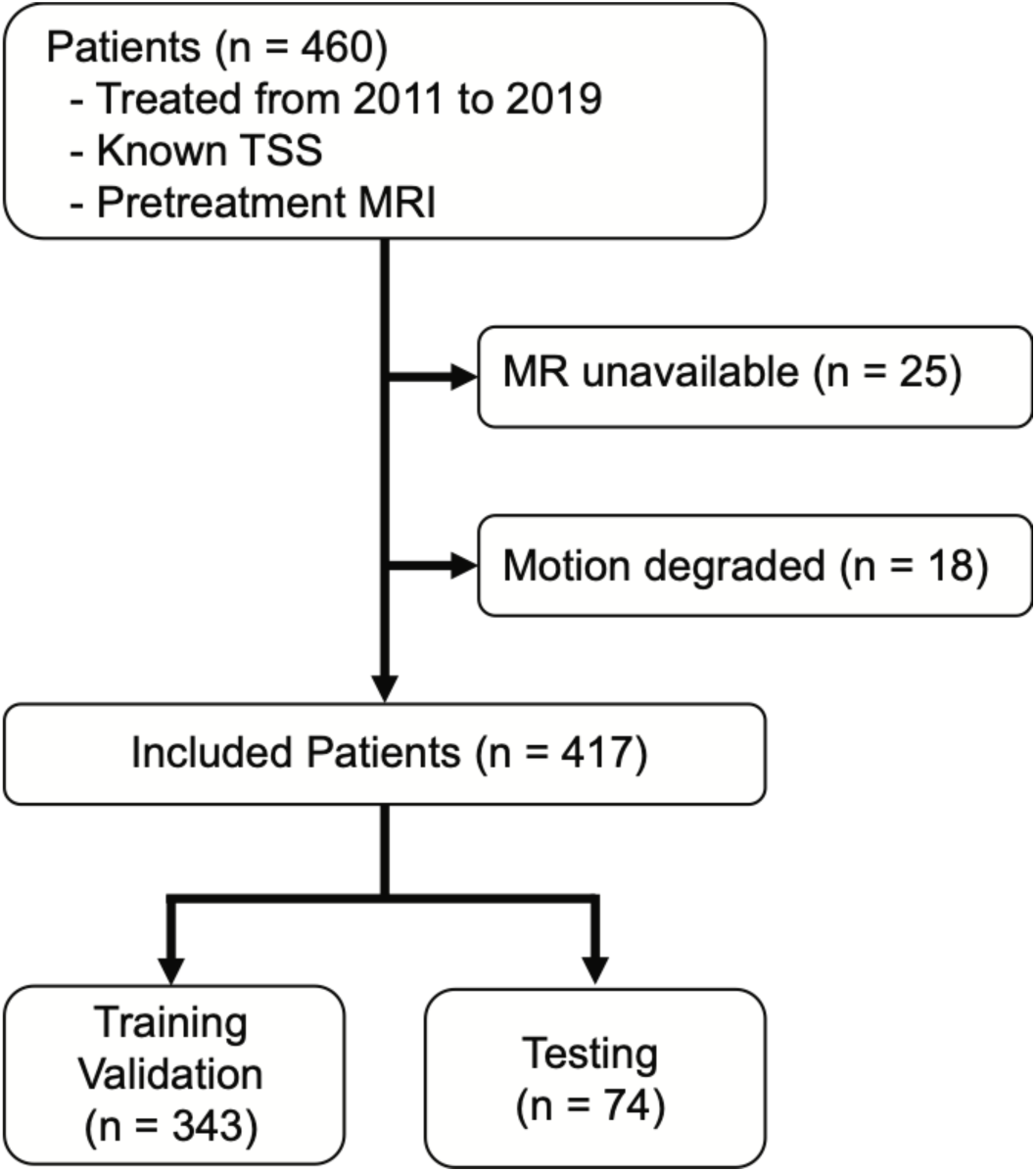
Patients were included based on clinical criteria. Patient flowchart illustrating inclusion criteria for this study.

### 3.2. DWI-FLAIR Mismatch Assessments

For each patient in the evaluation sets, three neuroradiologists independently assessed mismatch between DWI and FLAIR series. These labels served as a proxy for TSS, as mismatch indicates that a stroke occurred recently enough such that there are regions that have experienced ischemia (visible on DWI) but are not yet infarcted (visible on FLAIR). Radiologists performed these assessments on workstations within the same facility, and they were blinded to model classifications and EHR data. Final assessments were determined by majority vote among the three experts.

### 3.3. Model Evaluation

To train models, the development sets were split into five folds for cross-validated hyperparameter tuning, and the chosen parameters were used to train a model on the entire development set. Training was run in replication across ten random seeds. These trained models were run on the evaluation data, and metrics were computed and aggregated to generate confidence intervals. Metrics included sensitivity, specificity, accuracy, and receiving-operator characteristic area under the curve (AUC). The AUC analysis threshold was determined utilizing Youden’s Index on the training data. These statistics were compared to those of the majority radiologist classification. Additionally, Fleiss’ kappa was calculated to measure the level of agreement among the three radiologist assessments. We also report the inter-label agreement between the clinically recorded TSS and the DWI-FLAIR mismatch.

To evaluate the clinical utility of the machine learning and deep learning algorithms, we conducted the following experiments: 1) training and testing on data from the same institution, 2) training on one institution’s data and testing on the other, 3) training on data from both institutions.^21^ We report the performance on both internal and external evaluation sets.

### 3.4. Deep Learning Model

Following image pre-processing, the deep learning (DL) model utilized DWI, ADC, and FLAIR volumes. Model input encompassed three corresponding MRI slices, one from each series, of a single hemisphere of the brain. We designed a multi-slice model that utilizes weight sharing to extract neighboring slices’ spatial information. Image series were stacked as channels and fed into a shared convolutional layer and ResBlock. Intermediate features of neighboring slices were grouped and fed through five individual weight-sharing neighborhood subnetworks based on a ResNet34 backbone. The outputs were fed into a trainable softmax layer to fuse the features across subnetworks, enabling the model to learn the importance of certain subnetworks over others. Channel and spatial attention modules were attached at the last two ResBlocks to extract multi-scale features.^20^ Outputs were then fused with backbone features and fed through a fully connected layer to generate a patient-level TSS classification. A schematic of our model is illustrated in Figure 2. We adopted our previously published transfer learning schema to the model for the training process.^16^ The model was trained for 100 epochs with early stopping if validation AUC did not improve for five epochs. Binary cross-entropy was used as the loss function, with the Adam optimizer and weight decay, a learning rate of 0.0001, and a batch size of 12.

**Figure 2:**
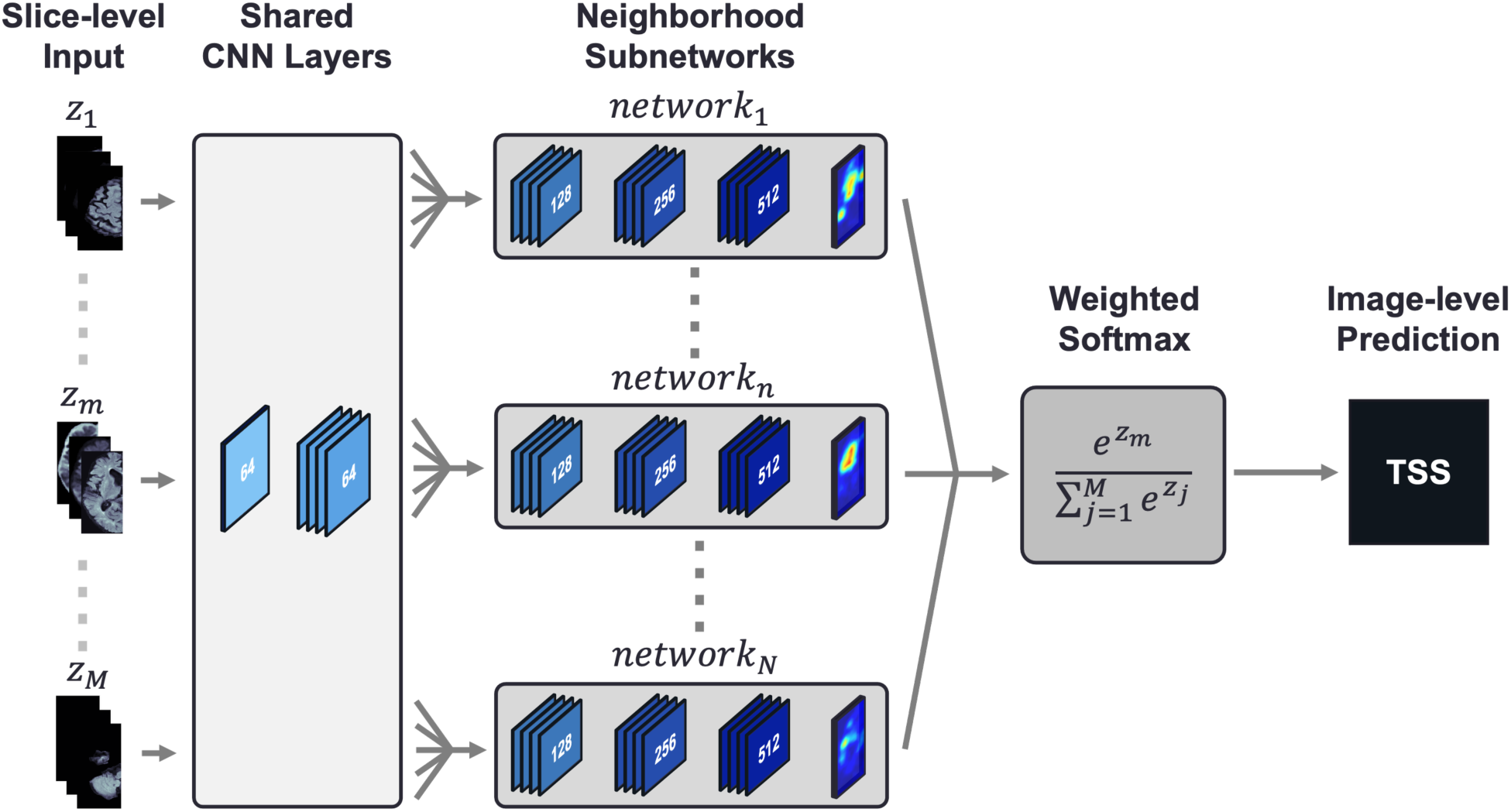
Convolutional neural network architecture with shared weights used to classify time since stroke (TSS) The deep learning architecture used DWI, FLAIR, and ADC series as input. The model split the volume into slices *z*_1_, …, *z*_*M*_ and stacks the image series as channels. Each slice *z*_*m*_ was fed into a shared set of convolutional layers. Intermediate output features from groups of adjacent slices were then propagated through five neighborhood subnetworks *network*_1_, …, *network*_*N*_, where weights are shared among the slice neighborhoods. Each subnetwork contained convolutional ResBlocks as well as convolutional attention modules to assist the model with localization. The resulting outputs from each subnetwork are aggregated using a weighted softmax function to generate a TSS classification for the image.

### 3.5. Comparison Model

A previously published radiomics machine learning (ML)^12^ method was also evaluated. The ML method began with infarct segmentation via normalized absolute thresholding. Regions of interest (ROI) were used as the basis for radiomic feature extraction, using DWI and FLAIR series and a FLAIR-ADC ratio map. These features were subjected to univariate *t*-tests to select the most informative features that were used in random forest, support vector machine, and logistic regression models.

### 3.6. Deep Learning Visualizations

We implemented three visualization methods used for model interpretability: occlusion sensitivity, class activation maps (CAMs), and integrated gradients. Each method provides unique feature importance maps for a given input. Occlusion sensitivity involves perturbing patches of input images and calculating the effect each perturbation has on the target class prediction.^22^ To generate CAMs, an activation map is computed using the output from the last convolutional layer of the network; this serves to identify regions of the image that provide the greatest discrimination for the correct label. Finally, outputs are backpropagated through the network to create pixelwise maps of network gradients for individual input images. We also visualize the class activation map (CAM) and gradients generated via guided backpropagation.^23^

### 3.7. Ethical Standards

This study conforms with World Medical Association Declaration of Helsinki. It was approved by the UCLA Medical Institutional Review Board #3 (MIRB3) under IRB#18-000329 “A Machine Learning Approach to Classifying Time Since Stroke using Medical Imaging”. Patient records were collected in accordance with IRB approval and HIPAA compliance standards. Informed consent was waived under Exemption 4 for retrospective data.

### 3.8. Data Availability Statement

The datasets presented in this article are not readily available due to protection of patient privacy. We are willing to validate other models internally on our data as part of collaborations. Program code and derived data will be made available at https://github.com/zhanghaoyue/stroke_tss_DL upon publication.

## 4. Results

Our study utilized two datasets. Of the internal set, 222 patients had a TSS under 4.5 hours, with the remaining 195 patients had a TSS over 4.5 hours. For external evaluation, we utilized a dataset totaling 355 patients, of which 182 underwent MRI within 4.5 hours of onset and 173 after 4.5 hours of onset.^12^ Clinical characteristics of these datasets are summarized in Table 1.

**Table 1:**
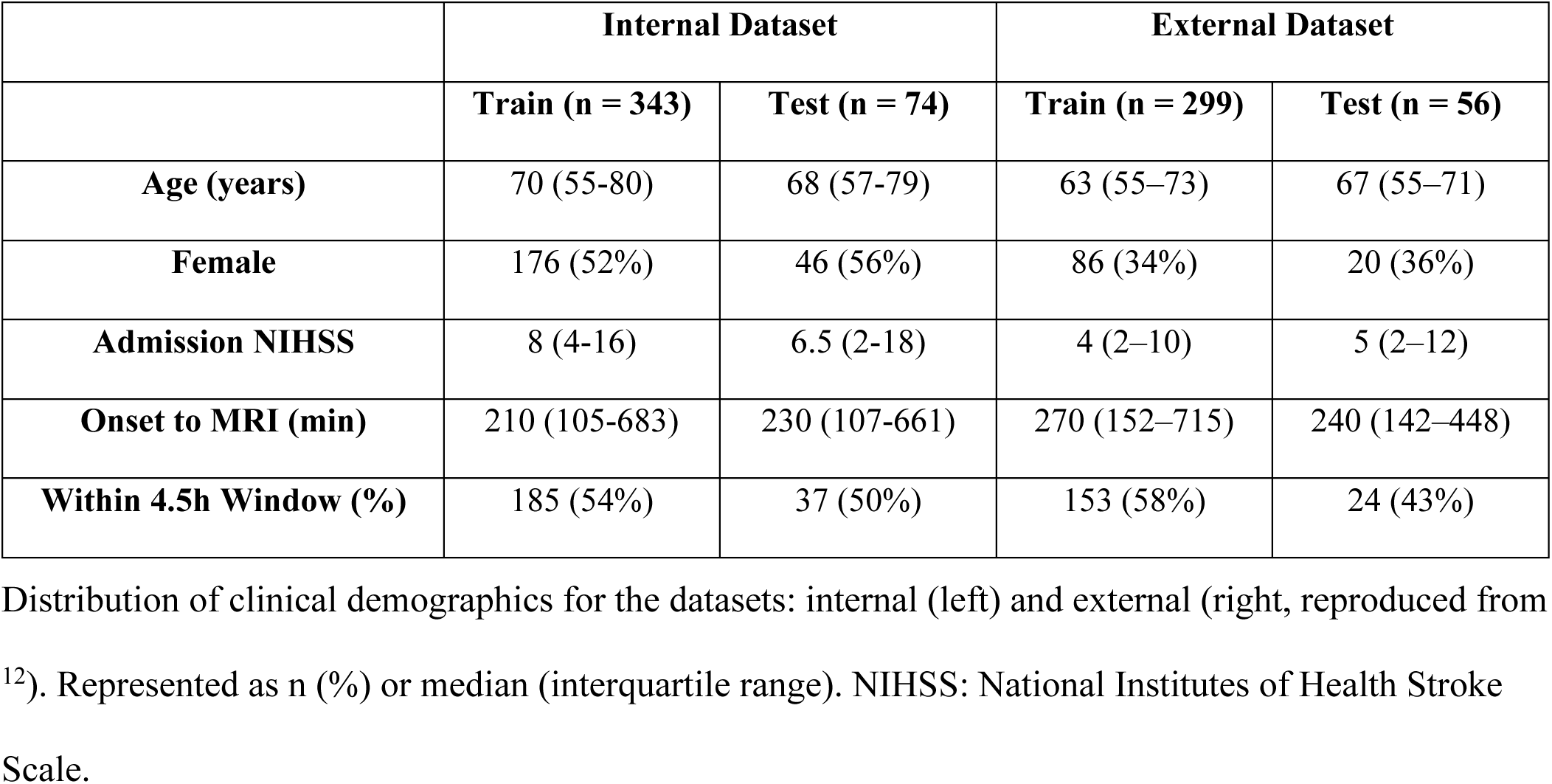
Patient Characteristics.

### 4.1. DWI-FLAIR Mismatch Assessments

Among the 130 patients assessed from both datasets, 37.8% (28/74) and 55% (31/54) of patients were found to have DWI-FLAIR mismatch in the internal and external evaluation sets, respectively. Inter-reader agreement among the radiologists as pairs and collectively are summarized in Table 2. Fleiss’ was 0.460 for the internal dataset and 0.575 for the external dataset, which are both typically classified as a moderate level of agreement. Performance of the human readers, compared to time clock assessment, is illustrated in Table 3 for the internal and external datasets. The majority radiologist assessment of mismatch for the internal evaluation set, when compared to the EHR-derived TSS, had low sensitivity (0.622) with high specificity (0.865). The aggregate assessment achieved higher accuracy (0.743) compared to the average accuracy of any individual radiologist (0.658). The mismatch assessments for the external evaluation set had higher sensitivity (0.743) while maintaining a high specificity (0.800).

**Table 2:** Inter-rater Agreement for the Internal and External Datasets.

| Site | Rad | Agreement ($\kappa$) |
| --- | --- | --- |
| Internal | Rad 1 – Rad 2 | 0.3677 |
|  | Rad 1 – Rad 3 | 0.5264 |
|  | Rad 2 – Rad 3 | 0.4879 |
|  | **All Radiologists** | **0.4600** |
|  | **Agg – TSS** | **0.4430** |
| External | Rad 1 – Rad 2 | 0.5893 |
|  | Rad 1 – Rad 3 | 0.6306 |
|  | Rad 2 – Rad 3 | 0.5086 |
|  | **All Radiologists** | **0.5755** |
|  | **Agg – TSS** | **0.5208** |

Calculated using Cohen's kappa, except for All Radiologists, which is computed Fleiss' kappa.

**Table 3:** Radiologist Performance Metrics.

| Site | Reader | Mismatch<br>Positive | Accuracy | Sensitivity | Specificity |
| --- | --- | --- | --- | --- | --- |
| Internal<br>(n=74) | Rad 1 | 38 | 0.608 | 0.568 | 0.649 |
|  | Rad 2 | 19 | 0.676 | 0.432 | 0.919 |
|  | Rad 3 | 28 | 0.689 | 0.541 | 0.838 |
|  | **Agg** | **28** | **0.743** | **0.622** | **0.865** |
| External<br>(n=56) | Rad 1 | 31 | 0.691 | 0.686 | 0.700 |
|  | Rad 2 | 35 | 0.836 | 0.857 | 0.800 |
|  | Rad 3 | 24 | 0.636 | 0.543 | 0.750 |
|  | **Agg** | **31** | **0.764** | **0.743** | **0.800** |

Performance metrics for individual and aggregate radiologist assessments for the internal and external datasets. Rad: Individual Radiologist. Agg: Aggregate reading by radiologists.

### 4.2. TSS Classification Models

The performance results of the DL and ML methods trained on the internal, external, and combination training sets, are summarized in Table 4. As a result of the thresholding technique applied by the ML method, 204 patients out of 417 patients from the internal dataset had an extracted ROI, and 343 out of 355 patients from the external dataset had an extracted ROI. Additionally, the ML model selected different radiomics features depending on the dataset. In applying univariate *t-*tests to 89 radiomics features, 37 features were selected for the internal training set and 35 were selected for the external training set with only seven features overlapping. When compared to the radiologist assessments, both the ML and DL model had higher sensitivity, though lower specificity. The average rate of agreement between the DL predictions and radiologist assessments was 0.411 (0.01), indicating a similar level of agreement as among the radiologists for the internal evaluation set.

**Table 4:** Performance Metrics.

|  | Train Set | Test Set | AUC | Accuracy | Sensitivity | Specificity |
| --- | --- | --- | --- | --- | --- | --- |
| **DL** | Internal<br>(n = 340) | Internal | 0.768 (0.03) | 0.726 (0.02) | 0.712 (0.08) | 0.741 (0.09) |
|  |  | External | 0.737 (0.03) | 0.724 (0.04) | 0.757 (0.04) | 0.679 (0.07) |
|  | External<br>(n = 299) | Internal | 0.732 (0.02) | 0.707 (0.03) | **0.716 (0.09)** | 0.687 (0.08) |
|  |  | External | 0.772 (0.02) | 0.767 (0.03) | **0.850 (0.08)** | 0.648 (0.09) |
|  | Both<br>(n = 639) | Internal | **0.840 (0.03)** | **0.789 (0.04)** | 0.777 (0.06) | **0.802 (0.07)** |
|  |  | External | **0.814 (0.01)** | **0.800 (0.04)** | 0.850 (0.08) | **0.727 (0.08)** |
| **ML** | Internal<br>(n = 164) | Internal | 0.730 (0.07) | 0.675 (0.07) | 0.405 (0.07) | 0.811 (0.08) |
|  |  | External | 0.680 (0.15) | 0.653 (0.10) | 0.714 (0.15) | 0.500 (0.13) |
|  | External<br>(n = 284) | Internal | 0.698 (0.08) | 0.625 (0.09) | 0.297 (0.08) | 0.865 (0.10) |
|  |  | External | 0.780 (0.05) | 0.735 (0.05) | 0.657 (0.05) | 0.800 (0.08) |
|  | Both<br>(n = 448) | Internal | 0.783 (0.03) | 0.750 (0.04) | 0.405 (0.03) | 0.892 (0.03) |
|  |  | External | 0.795 (0.03) | 0.735 (0.03) | 0.686 (0.03) | 0.750 (0.04) |

Performance metrics for the deep learning (DL) machine learning (ML)<sup>12</sup> models trained on the internal, external, and combination training sets, and tested on each separate test set. Statistics are reported as average (standard deviation).

The internally trained model achieved an AUC of 0.768 (0.03), with an accuracy of 0.726 (0.02), a sensitivity of 0.712 (0.08) and a specificity of 0.741 (0.09). On the external dataset, the model achieved an AUC of 0.737 (0.03), an accuracy of 0.724 (0.04), a sensitivity of 0.757 (0.04), and a specificity of 0.679 (0.07). When trained on the aggregate, performance on both evaluation sets improved, achieving an average AUC of 0.840 (0.03) on the internal dataset and 0.814 (0.01) on the external dataset. This aggregate model yielded an average accuracy of 0.794 (0.04), surpassing the accuracy of the aggregate neuroradiologist assessment.

### 4.3. External Evaluation

The impact of external training data on model AUC is summarized in Figure 3. The model achieved lower performance on the external evaluation set when no refinement is performed; however, the model achieved comparable performance for both evaluation sets when as few as 40 external samples were introduced into training, and better performance when 160 external patients were used. Intuitively, this corroborates the idea that deep learning algorithms achieve higher performance when trained on larger amounts of data and is illustrated in the second panel of Figure 3, where the performance on both cohorts did not improve with replacement of internal data with external data.

**Figure 3:**
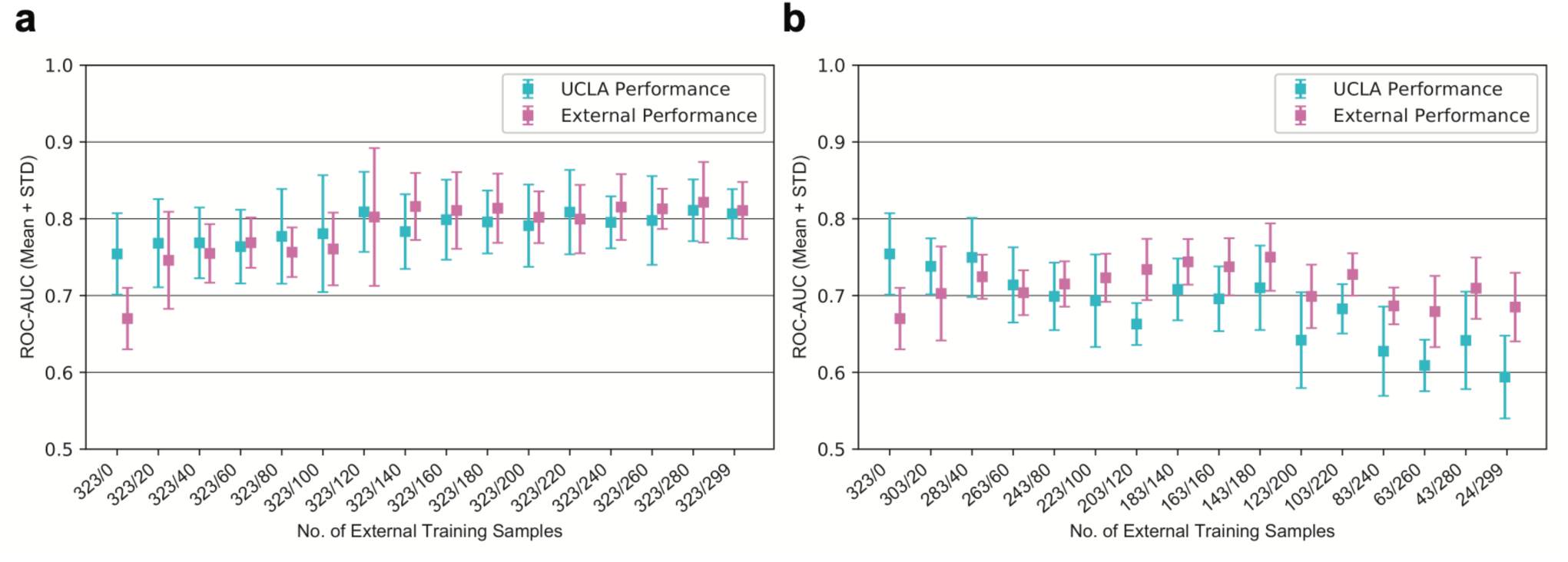
Performance of Models when Varying Training Data. Receiving-operator characteristic area under curve (AUC) of models with varying amounts of external training data, both when added to (**a**) or replacing (**b**) samples in the internal training set. Performance on both internal and external test sets are reported, in blue and pink, respectively, with 95% confidence intervals. Numbers on the x-axis indicate the number of internal/external samples used for training.

### 4.4. Deep Learning Visualizations

Visualizations were generated to reveal image regions on which the model focused. Four patients are shown in Figure 4. Figure 4A illustrates a case in which radiologist DWI-FLAIR assessment and TSS align with each other and the model prediction. The model does not solely focus on areas of high imaging signal, including the white matter hyperintensity seen on the FLAIR series, suggesting that our model localizes to lesions with other signal differences present. Figure 4B shows a case with a stroke onset time just under the 4.5-hour threshold that the neuroradiologists agreed contained no signal mismatch. In this instance, the model’s classification was outside the window. The gradients and CAM localize to the stroke lesion, while the occlusion method shows that areas outside the stroke volume were most salient to the prediction. Figure 4C shows a case just over the 4.5-hour threshold in which the radiologists were unanimous in identifying signal mismatch, despite the onset time being outside the window. Our model predicted this case to be within the treatment window. This discrepancy highlights that clock time may not encapsulate physiological state. Finally, Figure 4D shows a case well over the window for tPA. The radiologists agree that there is no mismatch, yet our model predicted that this case was within the window. The occlusion-based visualization shows that the model is unable to localize the stroke on either the ADC or FLAIR series. The class activation map (CAM) highlights that there is not a strong region of activation. Notably, the signal intensity of the stroke is relatively low, which may account for the model’s behavior. It is possible that changes to the preprocessing protocol may better distinguish the lesion and improve model performance for such cases.

**Figure 4:**
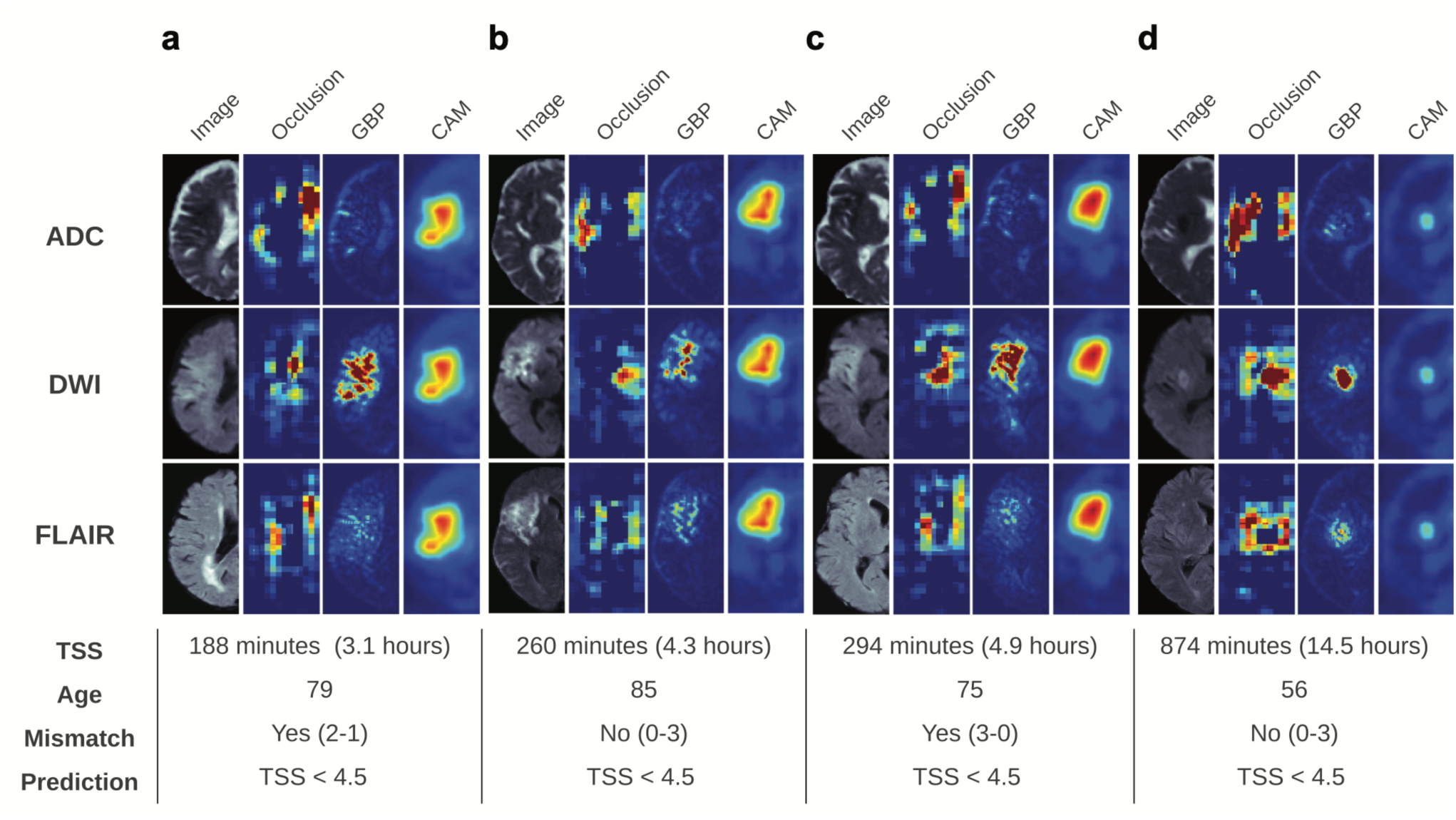
Deep learning visualizations demonstrate models focus on ischemic stroke regions. Deep learning algorithm visualizations for four patients (**a-d**), ordered with respect to time since stroke onset (TSS). For each patient, three visualizations were generated: occlusion, guided backpropagation (GBP), and class activation mapping (CAM). The table below lists the TSS, age, radiologist-assessed mismatch, and prediction yielded by the model.

## 5. Discussion

Our experiments yielded several findings. The radiologists’ readings for DWI-FLAIR mismatch were in moderate agreement. Our DL model achieved higher average performance than any of the ML models and higher sensitivity than the majority vote radiologist readings. The DL model was also able to generalize to an unseen external dataset. While there was a performance gap between internal and external evaluation sets, retraining the model with small amounts of external data improved classification performance.

The relationship between TSS and imaging features has been studied extensively; nevertheless, it remains unclear which signal patterns accurately capture the time course of ischemic tissue. DWI-FLAIR mismatch is one eligibility measure for thrombolysis in the most recent treatment guidelines. In our study, the inter-reader agreement for DWI-FLAIR mismatch aligns with that found in previous studies.^24,25^ Despite an average of 12 years’ experience among the neuroradiologists, variability among their assessments implies that a patient’s treatment options and therefore potential outcomes are reader dependent. Using TSS as the eligibility metric (“time clock”), the radiologist assessments identified 62% (23/37) of evaluation set patients who were within the 4.5-hour window of stroke onset. The DL model, by contrast, identified 76% (29/37) of patients within the window of eligibility. The lack of agreement among radiologists for the DWI-FLAIR mismatch assessments, along with the discrepancy between “tissue clock” and “time clock”, illustrate the need for more research into this relationship.

Our study reports the average performance of 10 replicates and evaluates two methods on the same datasets, revealing insight into the generalizability of these algorithms. When evaluated on external data, our model was able to achieve higher performance than the current state-of-the-art. This could be due to a few reasons: exclusion of potentially informative brain tissue when performing ROI extraction, and the bias introduced by statistical testing used for feature selection. Previous models, including the ML model evaluated in this study, have utilized segmentation models that identify the stroke region of interest from diffusion-weighted imaging.^12,26^ When compared to expert segmentation, performance of these methods has been moderate, primarily under-segmenting the stroke lesion. Moreover, these methods fail to incorporate penumbral regions that could inform vascular stroke progression status.^27^ In contrast, the DL model utilizes the ipsilateral brain hemisphere, thereby including information from both the ischemic core and the penumbral tissue outside diffusion-weighted lesions that may provide key insights into the tissue clock. Additionally, ROI extraction methods such as thresholding may exclude cases; utilizing brain hemispheres also keeps more cases that would not be able to be analyzed due to ROI generation process. For the ML model, the selection of statistically significant radiomics features may induce bias into the model that favors the training data; our DL model, in contrast, distills features from the entire input iteratively. The DL model also carries advantages over previous deep learning models^16^, likely as it uses attention modules to focus on pertinent channel and brain regions as well as the integration of information from neighboring slices. Despite these advantages, the DL model does have some drawbacks. The model has more input parameters than a standard radiomics-based ML model, requiring larger datasets and more computational time. This computation time is negligible for inference i.e., prediction, but should be considered for updating models when training. Additionally, there was still a performance gap for the DL model between the internal and external datasets, which motivated our external evaluation experiments. Aggregating the training datasets improves performance on both evaluation cohorts, indicating that DL classifiers improve synergistically when exposed to diverse training data.

Our study has several areas of potential improvement. While our dataset comprises the largest used for TSS classification from two cohorts, it cannot fully represent all patients seen in practice. Our preprocessing ideally minimizes dataset variation, but further analysis is needed to assess applicability to cohorts from other institutions. Second, we were only able to evaluate this model for a small set of patients for which the radiologists assessed mismatch. A common bottleneck when using machine learning for medical image tasks is that acquiring the label, e.g., having multiple neuroradiologists assess images for DWI-FLAIR mismatch, is labor-intensive and may not be feasible on a large scale. Third, TSS is not a perfect surrogate biomarker, as it does not always correlate to underlying tissue changes informing ischemia.^9^ Nonetheless, given the low inter-reader agreement of DWI-FLAIR mismatch, a TSS classification using an automated method may aid the radiologist in clinical decision-making.

Our proposed DL model allowed prediction of TSS based on MR images and achieved higher AUC than the ML model when external data was introduced, showing a more robust automated algorithm to determine stroke onset time. The results of this study indicate that a small amount of external data can improve generalized performance across patients from multiple institutions. These findings support the future study of implementation of a deep learning algorithm for clinical decision support in the setting of acute ischemic stroke treatment.

## Data Availability

The datasets presented in this article are not readily available due to protection of patient privacy. We are willing to validate other models internally on our data as part of collaborations. Program code and derived data (e.g., model weights) will be made available upon publication. Requests to access the datasets should be directed to the corresponding author.

## 7. Acknowledgments and Disclosure

We thank Dr. Hyunna Lee and Dr. Eun-Jae Lee at Asan Medical Center, University of Ulsan College of Medicine, for sharing their de-identified dataset with us and making their code publicly available, enabling us to reproduce their method for this study. The authors of this manuscript declare relationships with the following companies: Kambiz Nael, MD, serves as a consultant for Olea Medical. All other authors declare that the research was conducted in the absence of any commercial or financial relationships that could be construed as a potential conflict of interest.

